# Very low-calorie diet reduces hepatic steatosis and remodels circulating metabolite-microRNAs networks in metabolic dysfunction-associated steatotic liver disease: A pilot study

**DOI:** 10.64898/2026.09.01.26361664

**Authors:** Paroma Deb, Darin Bagar, Prashant Kumar, Leon Sun, Ethan Chen, Ravinder Reddy Gaddam, Lorrana Ferretto, Constance Shelsky, Antonio Sanchez, Himani Thakkar, Bhagirath Chaurasia, Ajit Vikram, Marcelo Lima de Gusmão Correia

## Abstract

Metabolic dysfunction-associated steatotic liver disease (MASLD) is a major cause of chronic liver disease, with weight loss as the pivotal therapeutic strategy. However, the metabolic and molecular adaptations underlying rapid weight loss remain incompletely defined. In this pilot study, women with obesity and MASLD but without diabetes consumed a very low-calorie diet (VLCD) for 8 weeks. Clinical parameters, hepatic steatosis measured by controlled attenuation parameter (CAP), circulating metabolites, and microRNAs (miRs) were assessed before and after the dietary intervention. Integrated correlation and hierarchical clustering analyses were performed to identify molecular networks associated with clinical improvement. VLCD was well tolerated, resulting in significant weight loss (∼11%) with ∼80% adherence. Significant improvements in metabolic parameters were observed, including fat mass, waist circumference, blood pressure, insulinemia, HOMA-IR, HbA1c, and triglycerides, with unchanged liver enzymes. Hepatic steatosis decreased markedly, as indicated by a reduction in CAP, while stiffness remained unchanged. Metabolomic profiling revealed elevated ketone bodies and broad reductions in amino acid levels, consistent with enhanced fatty acid oxidation and a catabolic metabolic state. Correlation analysis identified distinct metabolite signatures associated with hepatic steatosis, with changes in CAP positively associated with changes in amino acids and inversely associated with changes in ketone bodies and tricarboxylic acid cycle intermediates. Circulating miRs underwent selective rather than global remodeling, with only a limited subset showing strong associations with clinical parameters, including CAP and HOMA-IR. Specifically, VLCD altered the circulating levels of miR-148a-3p, miR-140-3p, miR-10b-5p, and miR-345-5p. Integration of metabolomic and miR datasets identified coordinated metabolite-miR modules involving glucose metabolism, branched-chain amino acid catabolism, mitochondrial metabolism, purine metabolism, microbial metabolites, and cellular redox pathways. These findings demonstrate that improvement in hepatic steatosis during VLCD-induced weight loss is accompanied by coordinated remodeling of circulating metabolite-miR networks. Integrated multi-omics analysis identifies candidate molecular signatures associated with metabolic adaptation and highlights circulating miR-metabolite modules as potential biomarkers of therapeutic response in MASLD.

## Introduction

Metabolic dysfunction-associated steatotic liver disease (MASLD) is a leading cause of chronic liver disease and a major contributor to liver-related morbidity and transplantation worldwide, with a global prevalence of ∼75% among people living with obesity.^1^ MASLD encompasses a spectrum from simple steatosis to metabolic dysfunction–associated steatohepatitis, which can progress to fibrosis, cirrhosis, and hepatocellular carcinoma. In addition to liver-related complications, MASLD is associated with increased risks of cardiovascular disease, chronic kidney disease, and sarcopenia, underscoring its systemic impact.^2^ Weight loss remains the cornerstone of MASLD management, with greater weight loss (e.g., ≥10%) associated with meaningful improvement in hepatic steatosis and MASLD.^3^ However, achieving and sustaining this degree of weight loss through conventional lifestyle modifications is challenging, and pharmacologic or surgical approaches are not suitable for all patients.^4^ Very low-calorie diets (VLCDs), which provide 800 kcal/day through nutritionally complete manufactured formulations, can induce rapid and substantial weight loss (>10% within 2–3 months) and have been shown to reduce hepatic steatosis in clinical studies.^5–7^ Despite these clinical benefits, the metabolic and molecular adaptations underlying rapid weight loss with VLCD remain incompletely defined. There is limited integration of circulating metabolites and microRNAs (miRs), which may provide insight into the systemic metabolic remodeling associated with hepatic improvement and serve as biomarkers of response. In this study, we investigated the effects of VLCD-induced weight loss in euglycemic women living with obesity and MASLD on hepatic steatosis, metabolic parameters, circulating metabolites, and miRs. We further examined the relationships between these molecular changes and clinical improvement to identify metabolic and regulatory signatures associated with reduced hepatic steatosis.

## Results

A total of 13 participants were recruited for the study; 3 participants dropped out due to noncompliance. A total of 10 participants, all female and aged 43 ± 3.7 years, completed the full study protocol (**Fig. 1**). Participants received VLCD for 8 weeks with ∼80% adherence. The intervention resulted in a significant reduction in weight, waist and neck circumferences, body fat percentage, blood pressure (systolic/diastolic), and HbA1c (**Table 1**). We did not observe any significant differences in fasting glucose, liver stiffness, alanine aminotransferase (ALT), aspartate aminotransferase (AST), and creatinine levels (**Table 1**). A marginal but statistically significant increase in uric acid was noted (**Table 1**). The controlled attenuation parameter (CAP), triglycerides (TAG), and the homeostatic model assessment of insulin resistance (HOMA-IR) showed significant decreases (**Fig. 2a-c**), indicating substantial changes in steatosis, lipid profile, and insulin resistance. These datasets show that the VLCD reduces body weight and liver steatosis.

**Fig. 1.**
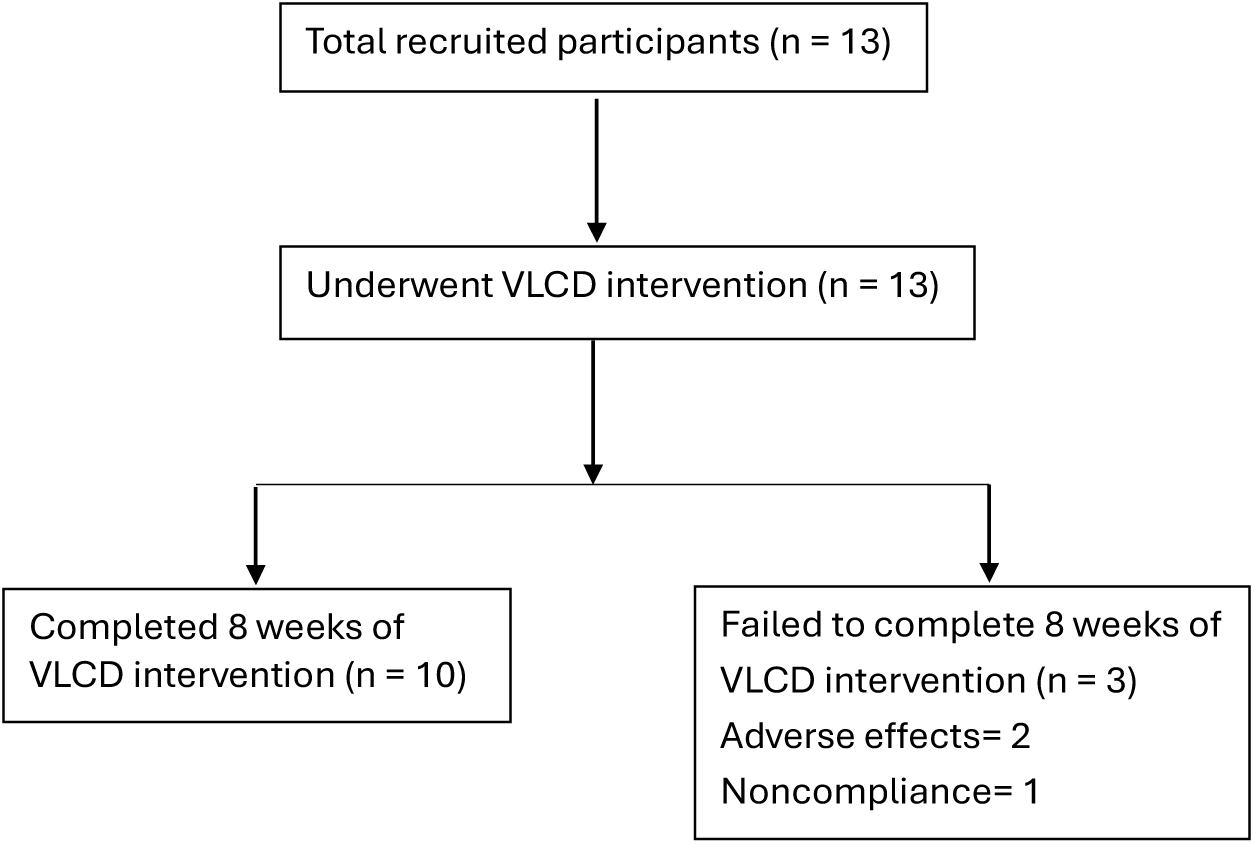
Flowchart illustration of the study participants.

**Fig. 2.**
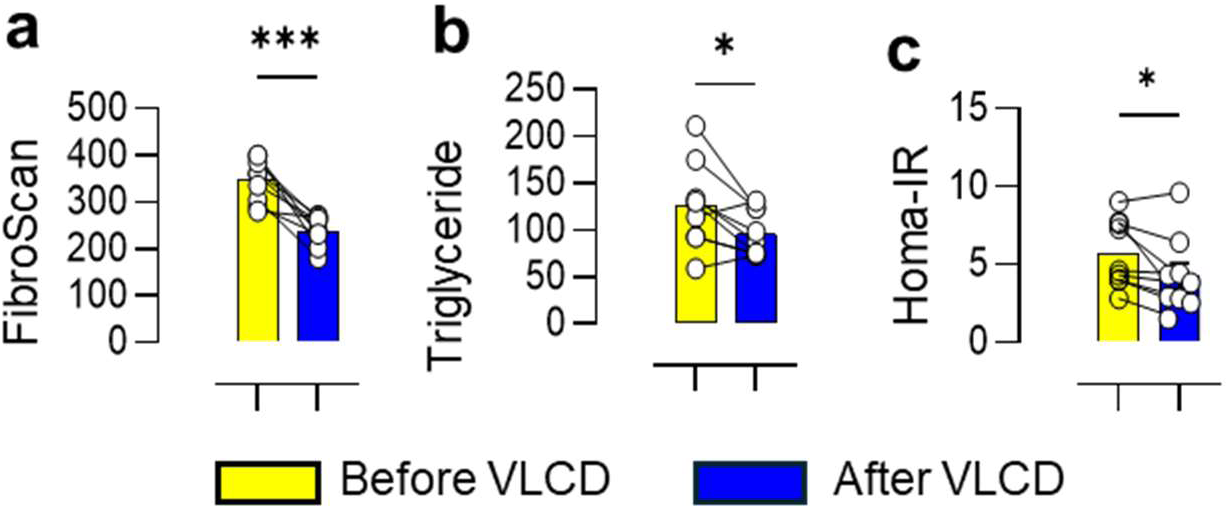
VLCD reduces hepatic steatosis and improves metabolic parameters in women with obesity. **a**) Liver steatosis measurement by elastography (FibroScan) before (yellow) and after (blue) 8 weeks of VLCD. **b**) Serum triglyceride levels before and after VLCD. **c**) Homeostatic Model Assessment for Insulin Resistance (HOMA-IR) before and after VLCD. Each dot represents an individual participant. Data are shown as mean ± SEM. Paired t-test was used; *p < 0.05, **p < 0.01, ***p < 0.001 *vs.* Before VLCD.

**Table 1.** Clinical characteristics in obese people pre- and post-VLCD intervention for 8 weeks.

|  |  |  |  |
| --- | --- | --- | --- |
|  | Before VLCD | After VLCD | P (Before VLCD vs. After VLCD) |
| Participants enrolled | 13 |  |  |
| Participants adhered to study protocol | 10 | 10 |  |
| Age (years) | 43 ± 3.71 |  |  |
| Sex | Female |  |  |
| Weight (kg) | 114.22 ± 3.58 | 101.5 ± 3.92 | <0.0001 |
| Body Mass Index (kg/m<sup>2</sup>) | 42.3 ± 1.45 | 38.3 ± 2.05 | 0.0015 |
| Body Fat Percentage (%) | 44.5 ± 0.73 | 42 ± 1.21 | 0.0039 |
| Waist Circumference (cm) | 130.13 ± 3.07 | 120.88 ± 3.95 | 0.0002 |
| Neck Circumference (cm) | 39.74 ± 0.80 | 37.62 ± 0.95 | <0.0001 |
| Systolic Blood Pressure (mmHg) | 126.73 ± 4.92 | 114.23 ± 3.06 | 0.0270 |
| Diastolic Blood Pressure (mmHg) | 79.58 ± 3.31 | 67.96 ± 2.30 | 0.0002 |
| Hemoglobin A1c (%) | 5.49 ± 0.13 | 5.17 ± 0.08 | 0.0009 |
| Fasting glucose (mg/dL) | 93.4 ± 3.77 | 91.9 ± 2.33 | 0.5282 |
| Liver Stiffness (kPa) | 7.37 ± 1.39 | 5.59 ± 0.70 | 0.1907 |
| Aspartate Aminotransferase (U/L) | 23.8 ± 3.08 | 25.9 ± 1.64 | 0.4283 |
| Alanine Aminotransferase (U/L) | 31.9 ± 5.47 | 36.5 ± 4.47 | 0.3936 |
| Creatinine (mg/dL) | 0.75 ± 0.04 | 0.74 ± 0.04 | 0.7208 |
| Uric Acid (mg/dL)* | 5.38 ± 0.29 | 5.85 ± 0.40<sup>^</sup> | 0.0480 |
| WBC (x10<sup>3</sup>/μL) | 8.32 ± 0.5 | 6.79 ± 0.61 | 0.0198 |
| Data are shown as mean ± SEM. *Uric Acid collected on visit 3. |  |  |  |

To characterize the systemic metabolic adaptations induced by VLCD, we profiled circulating metabolites and miRs before and after intervention. One participant was excluded due to hemolysis in the sample. VLCD induced a marked metabolic shift characterized by increased ketone body production and broad reductions in circulating amino acid levels, consistent with enhanced fatty acid oxidation and a catabolic state (**Fig. 3a**). To identify metabolites associated with clinical improvement, we performed pairwise correlation analyses between metabolites and clinical parameters. Correlation analysis identified distinct groups of metabolite-clinical parameter associations (**Fig. 3b**). Notably, the changes in several branched-chain amino acid (BCAA) catabolic intermediates, including α-ketoisovalerate (KIV), α-ketoisocaproate (KIC), and α-keto-methylvalerate (KMV), showed inverse correlations with changes in serum creatinine, whereas changes in methionine and anthranilate were positively associated with changes in platelet count (**Fig. 3b**). Changes in CAP were positively associated with changes in several amino acids (e.g., tyrosine, lysine, valine, tryptophan, cysteine) while negatively associated with changes in ketone bodies and tricarboxylic acid (TCA) cycle intermediates (e.g., 3-hydroxybutyrate, aconitate, citrate, glucose-6-phosphate, and erythrose) (**Fig. 3c**). The pairwise hierarchical clustering shows that in context of metabolites the HbA1c most closely resembled CAP among the measured clinical parameters (**Fig. 3c**). VLCD significantly altered several circulating miRs linked to insulin sensitivity and type 2 diabetes, including miR-148a-3p, miR-10b-5p, and miR-660-5p (**Fig. 4a**). Other miRs which are associated but not well-characterized in metabolic diseases, such as miR-30e-5p, miR-140-3p, 345-5p, and miR-10400-5p, were also significantly modulated (**Fig. 4b**). MiR-122 showed a trend toward increased levels following VLCD intervention (p = 0.06, **Fig. 4a**). To identify circulating miRs associated with clinical improvements, we performed pairwise correlation analysis between miRs and clinical parameters (**Fig. 4c**). Only a limited number of miR-clinical associations were observed, suggesting that metabolic improvement is accompanied by selective rather than global remodeling of circulating miRs. Specifically, changes in miR-23b and miR-320b were positively associated with changes in TAG, changes in miR-22b-3p were positively associated with changes in the insulin and HOMA-IR, whereas changes in miR-181a/b-5p were negatively associated with changes in platelet count (**Fig. 4c**). Changes in CAP were positively correlated with changes in miR-485-3p and miR-10400-5p while negatively associated with changes in miR-98-5p and miR-199b-5p (**Fig. 4d**). The pairwise hierarchical clustering in the context of miRs shows that HbA1c most closely resembled CAP among all measured clinical parameters (**Fig. 4d**).

**Fig. 3.**
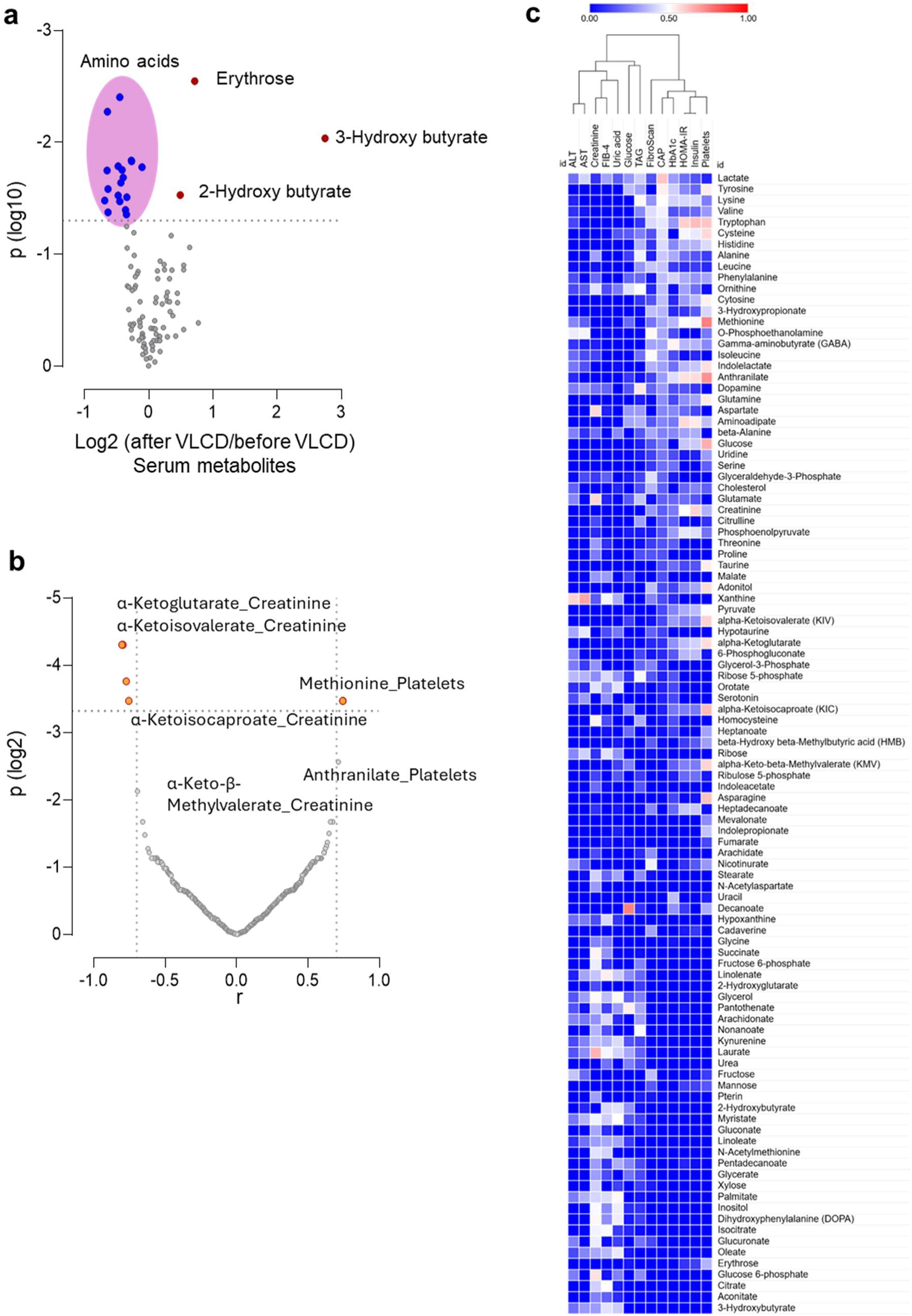
VLCD alters serum metabolites in women living with obesity. **a**) The volcano plot showing fold-change (log2) on the ‘x’ axis and significance (log10) on the ‘y’ axis of serum metabolites after 8 weeks of VLCD intervention. n = 9. **b**) The plot shows Pearson correlation coefficients (r) between changes in circulating metabolites and changes in clinical parameters on the ‘x’ axis and corresponding statistical significance (adjusted *P* value) on the ‘y’ axis. **c**) The heatmap showing Pearson correlation coefficients (r) between changes in circulating metabolites and changes in the clinical parameters. The metabolites are arranged according to their correlation with CAP. The columns are arranged based on pairwise hierarchical clustering.

**Fig. 4.**
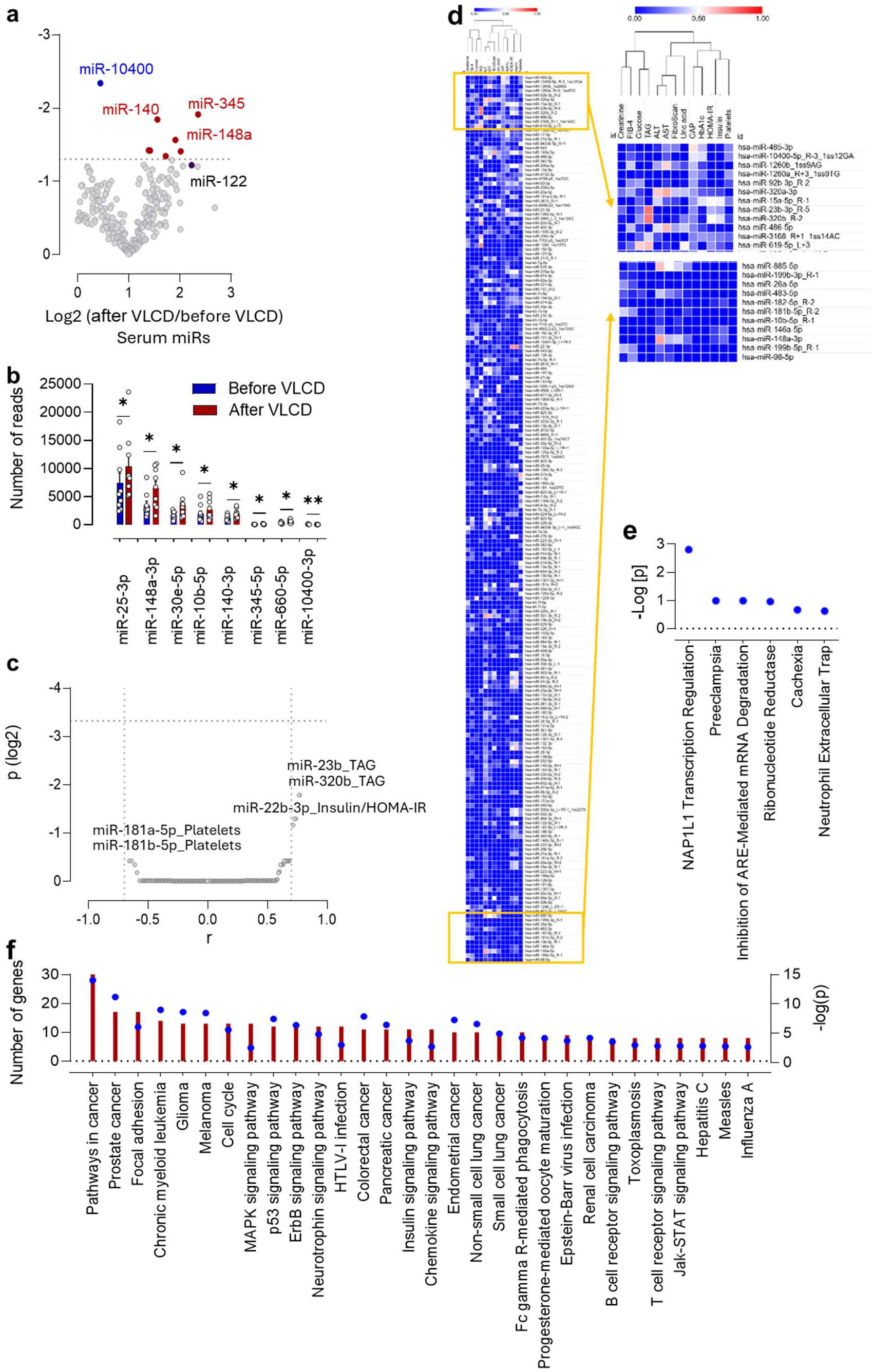
VLCD alters serum miR profiles in women living with obesity. **a**) The volcano plot shows fold-change on the ‘x’ axis (log2) and significance (log10) on the ‘y’ axis of serum miRs after VLCD. n = 9. **b**) The scatter plot shows the expression level of miRs that were significantly altered after VLCD. n = 9. **c**) The plot shows Pearson correlation coefficients (r) between changes in circulating miRs and changes in clinical parameters on the x-axis and corresponding statistical significance (adjusted *P* value) on the y-axis. **d**) The heatmap shows Pearson correlation coefficients (r) between changes in circulating miRs and changes in clinical parameters. The miRs are arranged according to their correlation with CAP. The columns are arranged based on pairwise hierarchical clustering. The zoomed-out section on the right shows the miRs that are associated with CAP. **e**) Signaling pathways identified via Ingenuity Pathway Analysis for differentially regulated miRs. **f**) miR-mRNA interaction analysis using miRNet 2.0 showing the top 30 dysregulated signaling pathways, with red bar indicating the gene count and the blue dots showing the statistical significance. Data in ‘b’ are shown as mean ± SEM. Paired t-test was used; *p < 0.05, **p < 0.01 *vs.* before VLCD.

Bioinformatic analyses of miRs identified pathways related to inflammation, cellular stress responses, and metabolic regulation (**Fig. 4e**). miR-mRNA interaction analysis using miRNet 2.0^8^ identified enriched KEGG (Kyoto Encyclopedia of Genes and Genomes) pathways related to cancer, cell adhesion, and cell cycle regulation, and inflammation (**Fig. 4f**).

To determine whether circulating miRs and metabolites underwent coordinated remodeling following VLCD, we performed pairwise correlation analyses of changes in all circulating miRs and metabolites. Although most associations were modest, several metabolite-miR pairs showed significant positive or negative correlations (**Fig. 5a**).

**Fig. 5.**
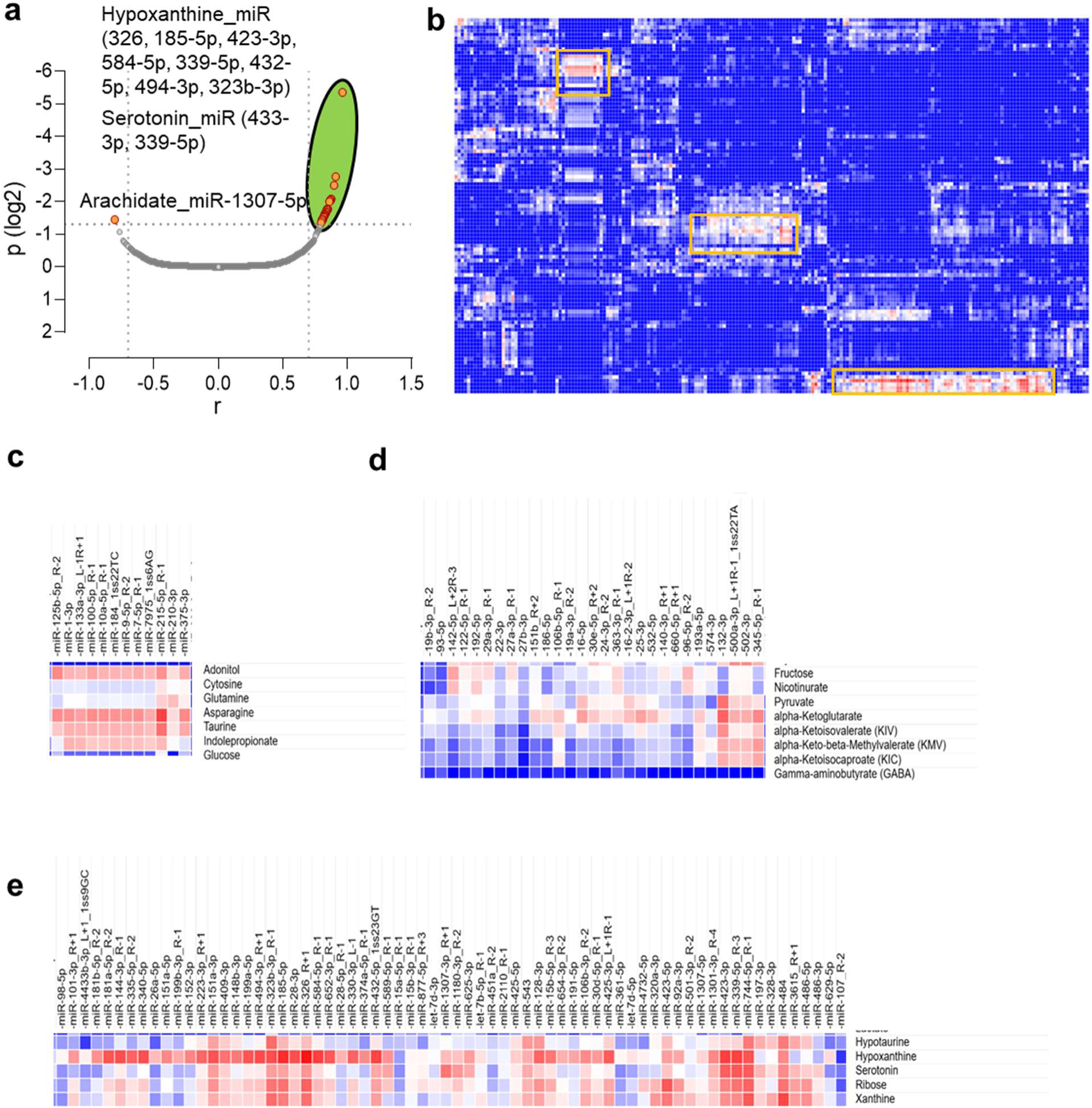
Coordinated remodeling of circulating miR-metabolite networks following VLCD. **a**) Plot showing Pearson correlation coefficients (r) between changes in circulating metabolites and changes in miRs on the ‘x’ axis and corresponding statistical significance (adjusted *P* value) on the ‘y’ axis. **b**) Heatmap showing Pearson correlation coefficients (r) between changes in circulating metabolites and changes in miRs. Rows and columns were hierarchically clustered to identify coordinated patterns of miR-metabolite associations. Yellow boxes indicate representative regions exhibiting the strongest coordinated associations. **c-e)** Enlarged views of the three representative miR-metabolite modules identified in panel b.

Hierarchical clustering revealed three major metabolite-miR clusters exhibiting coordinated correlation patterns (**Fig. 5b**). One miR-metabolite cluster included miRs (miR-125, miR-1, miR-133a, miR-100-5p, miR-10a-5p, miR-184, miR-9, let-7, miR-7975, miR-215, miR-210, and miR-375) that were strongly associated with metabolites linked to glucose homeostasis, amino acid metabolism, and microbial metabolism, including glucose, glutamine, asparagine, cytosine, adonitol, and indole-3-propionic acid (**Fig. 5c**). Another miR-metabolite cluster included miR-193b, miR-19b, miR-142-5p, miR-126-3p, miR-223, miR-29a, miR-30a/b, miR-27b, miR-215, miR-106b, miR-16, miR-25, miR-374a, miR-146b, miR-574, miR-502, and miR-345, which showed coordinated associations with metabolites, including fructose, pyruvate, α-ketoglutarate, branched-chain amino acid catabolic intermediates (KIV, KIC, and KMV), nicotinamide, and GABA (**Fig. 5d**). A third miR-metabolite cluster included miR-485-3p, miR-1260a/b, miR-92b-3p, miR-320a-3p, miR-766-3p, miR-145-5p, miR-451a, miR-21-5p, miR-361-5p, miR-30d-5p, miR-22-3p, miR-140-3p, miR-126-3p, and additional associated miRs, which showed coordinated associations with metabolites involved in purine metabolism and cellular redox pathways, including inosine, hypoxanthine, xanthine, ribose, sedoheptulose, and hypotaurine (**Fig. 5e**).

## Discussion

In this pilot study, we combined clinical phenotyping with metabolomic and circulating miR profiling to characterize the molecular adaptations accompanying VLCD-induced improvement in cardiometabolic parameters, insulin resistance, and hepatic steatosis in women with obesity complicated with MASLD. Hepatic steatosis, as reflected by CAP, decreased, whereas liver stiffness remained unchanged, consistent with previous short-term VLCD studies.^5–7^ Beyond confirming that VLCD markedly reduced hepatic steatosis and improved insulin sensitivity, our integrated analyses identified coordinated metabolite-miR networks associated with hepatic improvement, providing new insight into the systemic remodeling that accompanies rapid weight loss.

Metabolomic profiling revealed a shift toward increased fatty acid oxidation and reduced circulating amino acid levels, consistent with a catabolic state induced by caloric restriction. Importantly, pairwise correlation analysis of metabolites and clinical parameters revealed that VLCD-induced changes in BCAA catabolic intermediates were negatively correlated with changes in creatinine. miRs are key regulators of gene expression and play an important role in metabolic processes and disease.^9^ We demonstrate that VLCD-induced weight loss modulates circulating miR profiles, with several miRs previously linked to metabolic dysfunction showing dynamic changes following the intervention. Consistent with previous studies,^10,11,12^ VLCD significantly reduced circulating miR-148a-3p, miR-140-3p, and miR-345-5p. Moreover, miR-660-5p was observed with a higher expression level in children with metabolic-associated fatty liver disease,^13^ found to be downregulated in our study following VLCD intervention.

These miRs have previously been linked to metabolic dysfunction and hepatic steatosis, and KEGG analysis associated them with MAPK, insulin, and chemokine signaling pathways. We noted decreased levels of circulating miR-10400-5p following VLCD intervention. Several other circulating miRs such as miR-486-5p, miR-92a-3p, and miR-21-5p that are associated with obesity and metabolic dysfunction^10,12^ were also identified to be impacted by the VLCD intervention. Although circulating miR-122 is commonly elevated in obesity and MASLD,^14–17^ rapid caloric restriction may induce transient hepatic remodeling and lipid mobilization that alters miR-122 release independently of overall steatosis burden.

An important finding of this study is the integration of miR and metabolite data, revealing coordinated associations between miRs and metabolites. Hierarchical clustering demonstrated that metabolites involved in glucose utilization, amino acid metabolism, BCAA catabolism, mitochondrial metabolism, purine metabolism, and redox homeostasis clustered with distinct groups of circulating miRs. These observations suggest that VLCD-induced metabolic remodeling is organized into coordinated regulatory networks rather than occurring as independent biochemical changes.

Although correlation analysis cannot establish causality, these modules provide candidate pathways for future mechanistic investigation and may represent integrated molecular signatures of response to diverse dietary interventions.

We recognize that the small sample size and the inclusion of only female participants may limit the generalizability of the present study’s findings. In addition, the short duration of intervention limits assessment of long-term effects, particularly on fibrosis. Moreover, the absence of liver histology precludes direct evaluation of histological changes. The correlation-based analyses do not establish causality, and the observed miR-metabolite associations require validation in independent cohorts. Nevertheless, strict control of experimental and safety conditions, and its longitudinal design are methodological strengths of this pilot study.

In summary, our findings extend prior studies that have examined VLCD or VLCKD (very low-calorie ketogenic diet) effects on extracellular vesicles and oxidative stress responses^18, 19^ by providing an integrated view of circulating metabolites and miRs. Our results support VLCD as an effective short-term intervention to reduce hepatic steatosis and improve metabolic health. The coordinated metabolite-miR clusters provide a framework for future mechanistic studies and may serve as biomarkers for monitoring therapeutic response during dietary interventions in MASLD. Future studies with larger cohorts and longitudinal follow-up are needed to define the mechanistic roles and predictive utility of these molecular signatures.

## Methods

### Participants

The study population consisted of female participants aged ≥18 and <70 years with a body mass index (BMI) ≥30 and <50 kg/m². Eligible participants were required to test negative for viral hepatitis C (anti-hepatitis C antibodies) and autoimmune hepatitis (anti-smooth muscle antibodies). Exclusion criteria included type 1/type 2 diabetes mellitus, heart failure, myocardial infarction within the past 6 months, unstable angina, chronic kidney disease (eGFR <30 mL/min/1.73 m²), chronic obstructive pulmonary disease requiring oxygen supplementation, coexisting or end-stage liver disease, severe or uncontrolled psychiatric disorders (including eating disorders), gout, history of uric acid nephrolithiasis, porphyria, pregnancy or breastfeeding, active or prior cholecystitis without cholecystectomy, uncontrolled thyroid disease (TSH ≥10 mcIU/mL), excessive alcohol use (AUDIT-C score ≥3 for women), and use of medications including warfarin, lithium, or chronic prednisone (≥20 mg/day). Participants without elastography (FibroScan) within the prior 12 months were excluded. Individuals with F0 and S0 on elastography were excluded. Eligibility assessment included urine pregnancy testing (for women of childbearing potential), liver function tests (ALT, AST) if not available within 12 months, and elastography assessment. All participants completed the AUDIT-C questionnaire. Written informed consent was obtained before enrollment. All participants were recruited from the Weight Management Clinic and the Clinical Research Unit at the University of Iowa Health Care. The study was conducted in accordance with the Declaration of Helsinki and approved by the local Institutional Review Board.

### Study design

This pilot clinical study was a longitudinal, non-randomized, open-label trial designed to evaluate the effects of an 8-week VLCD intervention in individuals with obesity and MASLD. The study was approved by the University of Iowa Institutional Review Board (IRB no. 202008444). Written informed consent was obtained from each participant. The study consisted of two phases: baseline assessment and intervention. Eligible participants attended a baseline (week 0) visit at the University of Iowa Hospitals & Clinics Clinical Research Unit after a 12-hour fast. Eligibility criteria were confirmed, and a detailed clinical history and physical examination were performed. Alcohol use was assessed using the AUDIT-C questionnaire. Vital signs were recorded, and blood was collected for analysis of miRs, metabolomics, complete blood count, sodium, potassium, creatinine, uric acid, AST, ALT, glucose, triglycerides, insulin, and HbA1c. A urine pregnancy test was performed for women of reproductive age. Participants received standardized counseling on the VLCD protocol. Participants underwent an 8-week VLCD under the direct supervision of the principal investigator. Optifast® meal replacements were dispensed to participants at weeks 0, 2, and 4 at the Clinical Research Unit of the UIHC. At week 2, participants returned for a safety evaluation, including assessments of vital signs, sodium, potassium, and creatinine, as well as an adverse event assessment. At week 4, participants underwent repeat evaluation including assessment of sodium, potassium, creatinine, AST, ALT, and adverse event monitoring. At week 8, participants returned after a 12-hour fast for final assessments, including collection of blood for complete blood count, sodium, potassium, creatinine, AST, ALT, glucose, triglycerides, insulin, HbA1c, miRs, and metabolomics. Post-intervention elastography was performed. Participants also received counseling on transitioning from VLCD to a low-calorie (approximately 1200 kcal/day), low-fat diet.

Dietary adherence and body weight were assessed via telephone follow-up. Participants were instructed to record their body weight weekly. Body fat percentage was measured by bioimpedance, and neck and waist circumference were measured at each study visit.

### miR profiling

Total RNA was extracted from human serum samples using Qiazol/Trizol according to the manufacturer’s protocol. The quantity and purity of total RNA were measured using NanoDrop One Spectrophotometers (ThermoFisher Scientific, USA). LC Science (Houston, TX) performed the miR profiling as described.^20^ Briefly, 1 µg of RNA was used to construct the small RNA library, and quality control was performed using an Agilent Technologies 2100 Bioanalyzer High Sensitivity DNA Chip. Single-end (50 bp) sequencing was performed on the Illumina HiSeq 2500. Raw reads were processed using an in-house program, ACGT101-miR, to remove adapter dimers, junk, low-complexity sequences, common RNA families (rRNA, tRNA, snRNA, snoRNA), and repeats. To identify known and novel miRs, unique sequences with 18∼26 nucleotides were mapped to human-specific precursors in miRbase 22.0 by BLAST search. The unique sequences mapped to mature miRs in the hairpin arms were identified as known miRs, and sequences mapped to the opposite arm of the hairpin were novel 5p- or 3p- derived miRs. Modified global normalization is used to correct copy numbers among different samples.

### Metabolite profiling

Fresh serum samples were obtained by centrifugation of the coagulated blood and stored at −80°C. The samples were submitted to the Metabolomics core at the University of Iowa for analysis using a standard protocol. Briefly, the samples were diluted in ice-cold extraction solvent supplemented with internal standards. Serum extraction mixtures were vortexed for 10 minutes at room temperature and rotated for 1 hour at −20 °C. All the extraction mixtures were then centrifuged at 21,000 × *g* for 10 minutes at 4 °C, and 150 µL of the cleared supernatant was transferred into autosampler vials and dried using a SpeedVac vacuum concentrator (Thermo Fisher Scientific). The dried metabolite extracts were reconstituted in 30 µL of methoxyamine hydrochloride, derivatized with 20 µL of N, O-bis(trimethylsilyl)trifluoroacetamide + 1% trimethylsilyl chloride (BSTFA + 1% TMCS), and transferred to the gas chromatograph-mass spectrometer (GC-MS) autosampler for analysis. Derivatized samples (1 µL) were injected into a Trace 1300 GC equipped with a TraceGold TG-5SilMS column (Thermo Fisher Scientific). Metabolite detection was performed using an ISQ 7000 single quadrupole MS (Thermo Fisher Scientific). Acquired GC-MS data were processed by Thermo Scientific TraceFinder 5.1 software, and metabolites were identified based on the University of Iowa Metabolomics Core facility standard in-house library. NOREVA was used to correct signal drift. Corrected peak intensities were normalized to the total signal per sample to account for variability in extraction, derivatization, and sample loading.

### Correlation and statistical analysis

To evaluate the relationships among circulating miRs, metabolites, and clinical measures, Pearson correlation analyses were performed in R. Data wrangling and construction of the correlation matrix were carried out using the tidyverse framework. For correlation analyses, changes in clinical parameters, metabolites, and miRs were calculated for each participant as the difference between post-intervention and baseline values. Correlations were evaluated for three sets of analyses: changes in metabolites versus changes in clinical variables, changes in miRs versus changes in clinical variables, and changes in metabolites versus changes in miRs. Corresponding P values were adjusted for multiple testing using the Benjamini–Hochberg false discovery rate correction. Clustered heatmaps were generated using the Broad Institute’s Morpheus Web application.

## Data Availability

All data produced in the present study are available upon reasonable request to the authors.

## Author contributions

MLGC supervised participant recruitment and blood collection. PD, HT, and RRG procured, stored, and processed blood samples for metabolomics and miRs profiling. PD, PK, BC, and AV performed the bioinformatic analysis. PD, AV, DB, and MLGC prepared the first draft of the manuscript. DB, LF, AS, and MLGC contributed to writing the clinical sections of the results section. LS and EC organized the data in Table 1, analyzed the clinical data, and contributed to the writing of the introduction. MLGC and AV conceptualized the study. AV interpreted and integrated metabolomic and miR findings and developed the scientific framework. AV and MLGC secured funding and provided overall scientific supervision of the project. All authors reviewed and approved the final version of the manuscript.

## Acknowledgments

This work was supported by grants from the Stead Family Funds and the Investment in Strategic Priorities (ISP) Funding (University of Iowa) to MLGC and the University of Iowa Startup fund to AV. AV was partly supported by the NIHR01HL167773. PD was partly supported by the FOEDRC Bridge-to-the-Cure fund. RRG was partly supported by the AHA postdoctoral award (828081) and Career Development Grant (23CDA1037711).

## Conflict of Interest

The authors declare no conflict of interest.

## Notes

### Competing Interest Statement

The authors have declared no competing interest.

### Clinical Trial

NCT04861571

### Author Declarations

IRB committee of The University of Iowa gave ethical approval for this work.

